# Home-based transcranial direct current stimulation (tDCS) in major depressive disorder: enhanced network synchronization with active relative to sham and deep learning-based predictors of remission

**DOI:** 10.1101/2024.06.10.24308593

**Authors:** Wenyi Xiao, Jijomon C. Moncy, Rachel D. Woodham, Sudhakar Selvaraj, Nahed Lajmi, Harriet Hobday, Gabrielle Sheehan, Ali-Reza Ghazi-Noori, Peter J. Lagerberg, Rodrigo Machado-Vieira, Jair C. Soares, Allan H. Young, Cynthia H.Y. Fu

**Author notes:** Authors for correspondence: Dr. Wenyi Xiao, University of East London, School of Psychology, Arthur Edwards Building, Water Lane, London E15 4LZ, UK, Professor CHY Fu, University of East London, School of Psychology, Arthur Edwards Building, Water Lane, London E15 4LZ, UK.

## Abstract

**Aim:** To investigate neural oscillatory networks in major depressive disorder (MDD), effects of home-based transcranial direct current stimulation (tDCS) treatment, and potential predictors of treatment remission.

**Methods:** In a randomised controlled trial (RCT) of home-based tDCS treatment, EEG data were acquired a subset: 21 MDD participants (16 women) (mean age 36.63 ± 9.71 years) in current depressive episode of moderate to severe severity (mean Hamilton Depression Rating Scale (HAMD) score 18.42 ± 1.80). Participants were randomised to either active (n=11) or sham tDCS (n=8). Treatment was home-based tDCS treatment for 10 weeks in a bifrontal montage (anode over left dorsolateral prefrontal cortex) consisting of 5 sessions per week for 3 weeks and 3 sessions per week for 7 weeks. Active tDCS was 2mA and sham tDCS 0mA with brief ramp up and ramp down period to mimic active stimulation. Each session was 30 minutes. Clinical remission was defined as HAMD score ≤ 7. Resting-state EEG data were acquired at baseline, prior to the start of treatment, and at 10-week end of treatment. EEG data were acquired using portable 4-channel EEG device (electrode positions: AF7, AF8, TP9, TP10). EEG band power was extracted for each electrode and functional connectivity phase synchronization by phase locking value (PLV). Deep learning was applied to baseline PLV features to identify predictors of treatment remission.

**Results:** Main effect of group was observed in gamma PLV in frontal and temporal regions, in which active tDCS treatment group showed higher connectivity as compared to sham group. In active treatment group, significant positive correlations between changes in delta, theta, alpha, and beta PLV and improvement in depression severity were observed. The highest treatment remission prediction was achieved by combining PLV features from theta, alpha, and beta: accuracy 71.94% (sensitivity 52.88%, specificity 83.06%).

**Conclusions:** Synchronized brain activity across large-scale networks as reflected in gamma PLV is a potential mechanism of active tDCS as compared to placebo-sham tDCS. Baseline resting-state EEG is a potential predictor of treatment remission. Home-based EEG measures are feasible and potentially useful predictors of clinical outcome.

## 1. Introduction

Major depressive disorder (MDD) is a common and debilitating mental health disorder, characterized by persistent feelings of a low mood or inability to experience pleasure that is associated with a diminished interest in daily activities and changes in neurovegetative symptoms (American Psychiatric Association & Association, 2013). MDD is a heterogenous disorder, in which the remission rate is around 30-40% following the initial medication trial and about 55-55% after a subsequent trial, and there can be a lengthy process of trial and error to identify the optimal treatment (Trivedi et al., 2006; Fu et al., 2019).

The non-invasive brain stimulation, transcranial direct current stimulation (tDCS), is emerging as a potential treatment option in MDD (Mutz et al., 2018, 2019; Woodham et al., 2021). tDCS involves the application of a low-level electric current to the scalp, typically between 0.5-2.0 mA, with the anode most commonly placed over the left dorsolateral prefrontal cortex (DLPFC) in MDD treatment studies (Mutz et al., 2018, 2019, Moffa et al., 2020). The current modulates neuronal activity and neural regions more widely involved in mood regulation beyond the directly targeted areas (Polania et al., 2011).

As tDCS is portable and has a strong safety profile, we developed a home-based protocol which has demonstrated significant efficacy in a multi-site randomised controlled trial (Woodham et al., 2022, 2023). To investigate the neurophysiological effects and potential predictors of treatment response, electroencephalography (EEG) data were acquired in a subset of participants. EEG is a promising method for identifying neurobiological predictors of treatment response. Measures of connectivity, asymmetry across hemispheres, and power in frontal and temporal electrodes are predictive of treatment response to antidepressant medication as well as to repetitive transcranial magnetic stimulation (rTMS) (Watts et al., 2022). Applying deep learning, treatment response to rTMS was predicted with accuracies over 90% (Adamson et al., 2022).

In the present study, we sought to investigate the effects of a home-based tDCS treatment in MDD and to identify potential predictors of clinical response. We employed a portable wireless EEG device equipped with 4 dry electrodes, known for its robust signal properties (Cannard et al., 2021; Krigolson et al., 2021). Participants underwent EEG recording in their own homes under real-time supervision via video conference. We analysed EEG metrics, including power and phase locking value (PLV). PLV evaluates the consistency of phase differences over time, making it useful for detecting phase synchronization even in low-amplitude signals. This offers a complementary perspective to power measurements, which quantify signal strength. We employed deep learning methods to explore whether baseline EEG measures could predict treatment remission.

## 2. Method

### 2.1 Participants

Ethical approval was obtained from South Central-Hampshire B Research Ethics Committee. All participants provided written informed consent electronically. The study was a double-blind, placebo-controlled, randomized, superiority, trial of home-based tDCS in MDD (Woodham et al., 2023). Participants were 18 years or above, having a MDD diagnosis and currently experiencing a major depressive episode without psychotic features, based on Diagnostic and Statistical Manual of Mental Disorders, Fifth Edition (DSM-5) (American Psychiatric Association & Association, 2013) criteria and determined by a structured assessment using Mini-International Neuropsychiatric Interview (MINI; Version 7.0.2) (Sheehan, 1998). All participants had at least a moderate severity of depressive symptoms, as measured by 17-item Hamilton Depression Rating Score (HAMD) of ≥ 16 (Hamilton, 1960).

In inclusion criteria, participants were medication-free, or were taking antidepressant medication for at least 6 weeks, or were in psychotherapy for at least 6 weeks prior to enrolment. Exclusion criteria included having a history of mania or psychosis, having a neurological disorder or a medical disorder that may mimic mood disorders, significant suicide risk, or any exclusion criteria for receiving tDCS. Participants were randomly assigned to receive either active tDCS treatment (n=11; 8 women; mean age 38.45 years, SD 9.67 years) or sham tDCS treatment (n=8; 8 women; mean age 34.16 years, SD 9.99 years) (Table 1). Further information regarding demographic and clinical differences between the treatment groups is available in the Supplementary Materials.

**Table 1.**
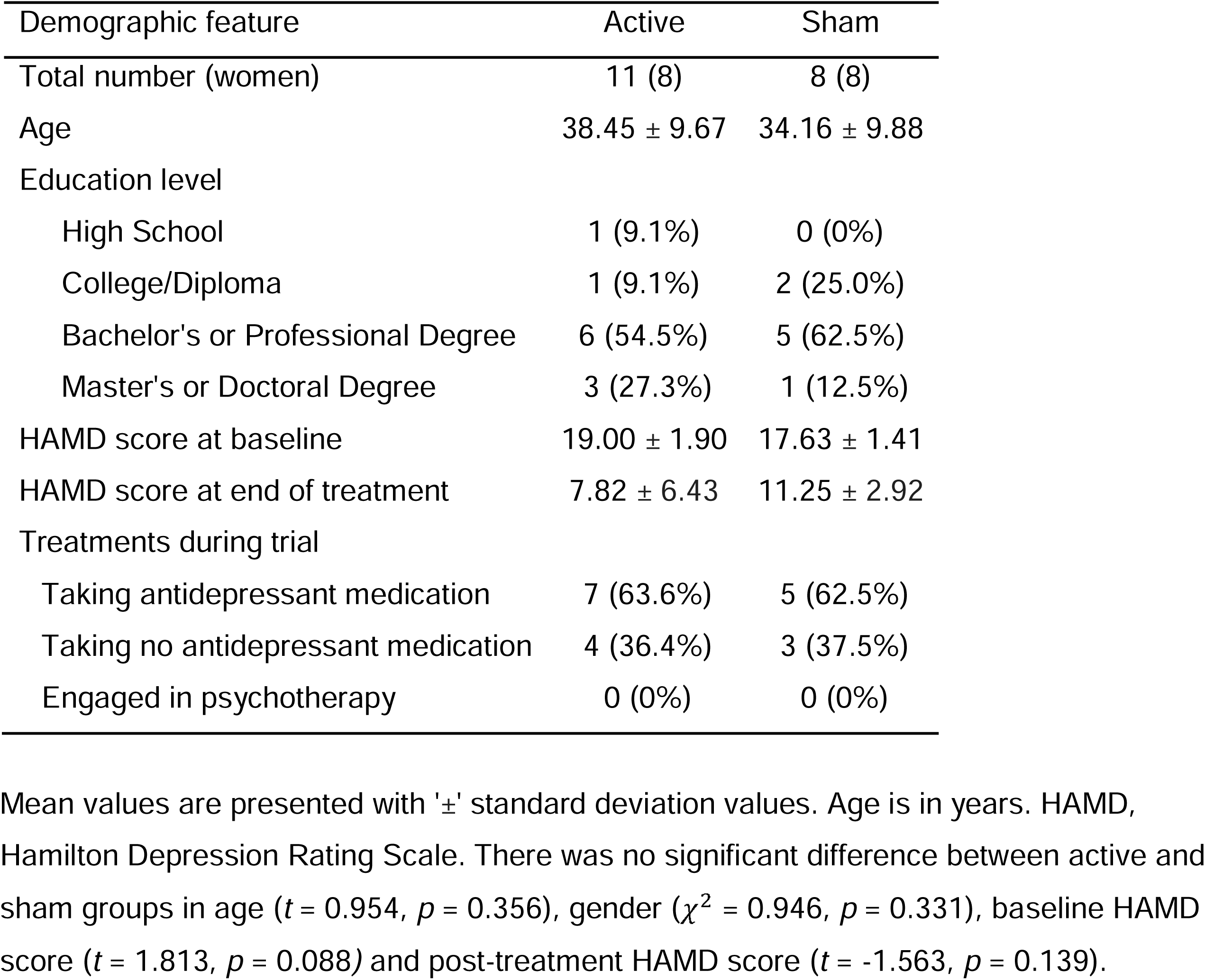
Demographic characteristics of participants

### 2.2 tDCS treatment protocol

Protocol consisted of a 10-week course of active or sham tDCS, which was self-administered by participants in their homes 5 times a week for 3 weeks and then 3 times a week for 7 weeks, for a total of 36 sessions. A member of the research team was present for the first session by Microsoft Teams video call. Bifrontal montage was applied with the anode positioned over left DLPFC (F3 position according to international 10/20 EEG system) and cathode over right DLPFC (F4 position). Each electrode was a 23cm^2^ conductive rubber electrode covered by saline soaked sponges. Active simulation was 2 mA for a duration of 30 minutes with a gradual ramp up over 120 seconds at the start and ramp down over 15 seconds at the end of each session. Sham stimulation had an initial ramp up from 0 to 1 mA over 30 seconds then ramp down to 0 mA over 15 seconds, which was repeated at end of the session to provide a tingling sensation that mimics active stimulation. Flow Neuroscience tDCS device was used for all participants. Clinical remission was defined as HAMD score of ≤ 7 at the 10-week end of the treatment.

### 2.3 Remote EEG acquisition and preprocessing

EEG data were collected at two time points: at baseline, prior to the start of treatment (baseline), and at the 10-week end of treatment (post-treatment). EEG data had been acquired in sub-sample of 21 MDD participants (18 women; mean age 37.10 years, SD 9.71 years) at baseline. However, post-treatment data were not available for one participant and was of consistently poor quality for another participant and was not included. Thus, data were available in 19 MDD participants (16 women; mean age 36.63 years, SD 9.71 years) at both timepoints (Table 1).

At each EEG acquisition session, a trained research team member provided real-time guidance via videoconference. Four 5-minute EEG recordings were conducted at each timepoint, with participants instructed to maintain a relaxed posture without movements. Resting state recordings were conducted in the following sequence: eyes closed, eyes open, eyes closed, and eyes open. In the present analysis, the two 5-minute resting state eyes-closed recordings at both timepoints were utilized.

EEG recordings were acquired using a portable, wireless MUSE device with 4 dry electrodes and sampling frequency of 256 Hz. Electrodes were positioned at AF7, AF8 (frontal sites), TP9, and TP10 (temporoparietal sites), with reference to the FPz electrode. The recorded EEG signals, saved in CSV format, included timestamps for each EEG sample, raw EEG signals from each electrode, and Horse Shoe Indicator (HSI) values for each electrode, indicating electrode connectivity quality. EEG signals were windowed into 10-seconds long windows without overlap. HSI values, ranges from 1 (excellent connectivity), 2 (medium connectivity), and 4 (poor connectivity), were available for each EEG sample for quality assessment. These values were averaged across samples within each window, and windows with an average HSI value of 2 or less were selected for further analysis. EEG signals from each electrode were filtered across six frequency bands: the full band (1-60 Hz), delta δ (1-4 Hz), theta θ (4-8 Hz), alpha α (8-12 Hz), beta (12-30 Hz), and gamma (30-60 Hz). Butterworth Infinite Impulse Response (IIR) band-pass filters of 5th order were utilized for filtering of the EEG signals.

### 2.4 Aggregation of rs-EEG measures

Resting state EEG metrics, including EEG band power atable nd phase locking value (PLV), were extracted. EEG band power was calculated for all four electrodes (AF7, AF8, TP9, TP10). PLV was computed for all possible electrode pairs (AF7-AF8, AF7-TP9, AF7-TP10, AF8-TP9, AF8-TP10, TP9-TP10). PLV evaluates the phase synchrony between two time-series signals (Hoke et al., 1989; Lachaux et al., 1999) and is commonly used to assess functional connectivity between EEG signals from different electrodes, revealing temporal relationships of neural signals irrespective of their amplitude. PLV is a statistical metric constrained within the range of 0 to 1. PLV value approaching 1 indicates high phase synchronization with minimum variation in phase difference across the EEG signals, while a value close to 0 suggests no phase synchronization. A total of 24 band power values and 36 PLVs were computed using two sets of 60 EEG measurements each at pre- and post-treatment, available for statistical analysis. Additional details are provided in the Supplementary Materials.

### 2.5 Statistical analyses

Statistical analyses were conducted on band power and PLV to explore potential associations between changes in EEG metrics and the severity of depression, as well as to evaluate the effects of active and sham tDCS treatment.

Pearson’s correlation was conducted to examine the relationship between changes in EEG measures and proportional change in HAMD scores over the 10-week treatment period across all participants. The proportional change in HAMD scores was determined by subtracting baseline score from post-treatment score and then dividing absolute difference by baseline score. Factorial analyses with and without proportional change in MADRS as covariate were used for between-group comparisons. To test whether EEG measures changed in response to treatment, a two-way ANOVA was performed for each EEG variable. The factors were: remission group (remission, non-remission) and time (baseline, post-treatment). 60 statistical tests were performed in total. Additional details are provided in the Supplementary Materials.

Coefficients were estimated in the R statistical environment (R Core Team, R. (2013)) using linear regression (*lm* built-in function). Post hoc tests (Tukey honestly significant difference (HSD)) were performed to assess significant effects. The statistical threshold was set at *p* < 0.05, with correction for multiple comparisons by controlling False Discovery Rate (FDR). A full description is in the supplementary materials.

### 2.6 Deep learning analysis

Participants were categorized into two groups based on their remission status following treatment. In the deep learning-based classification analysis, participants achieved remission were labelled as the positive class and non-remission as the negative class, with sensitivity representing remission and specificity representing non-remission. For predicting treatment response, PLV values were used and were extracted from different frequency bands of pre-treatment EEG. From each EEG frequency band, PLV feature vector with a dimension 6 were generated, representing each of the six electrode pairs. These six-dimensional feature vectors, both individually and through the concatenation of multiple EEG bands, were employed as inputs for deep learning models with varying parameters. This concatenation process at the feature level led to a linear increase in the feature dimension. To assess the effectiveness of different combinations of PLV features from individual bands and combinations of multiple bands, two distinct deep learning architectures were investigated. The first architecture was a fully connected perceptron deep learning structure, and the second employed a one-dimensional convolutional neural network (1DCNN) architecture. Considering combination of features from multiple EEG bands, the dimensionality varied as follows: 6 (single band), 12 (two-band combination), 18 (three-band combination), 24 (four-band combination), and 30 (combination of all bands).

For the fully connected perceptron deep learning network, a four-layer architecture was implemented, comprising layers with 32, 32, 16, and 1 perceptrons (output layer) for all input combinations, except in the case of the all-band combination where the feature size was 30. In this scenario, 64 perceptrons were employed in the first layer. The all-band combination encompassed all PLV features extracted from the delta to gamma bands. In the 1DCNN-based architecture, the initial fully connected layer of the perceptron network was replaced with a convolutional layer. This convolutional layer employed a kernel size of 3. To ensure kernel overlap, the number of filters used was determined by multiplying the input dimension by a multiplication factor 2/3. Following the convolutional layer, a MaxPooling1D layer with a pool size of 2 was integrated to downsample the feature maps, aiming to extract the most relevant features while reducing computational complexity. For single-band PLV features with an input dimension of 6, 4 filters with a kernel size of 3 were used. Similarly, for dimensions 12, 18, 24, and 30, 8, 12, 16, and 20 filters were employed respectively. The activation function ’relu’ was applied to all layers except the output layer, where ’sigmoid’ was utilized. To minimize the ’binary cross-entropy’ loss function, the ’adam’ optimizer was employed.

Due to the constrained size of the dataset and our emphasis on accurately evaluating model performance over computational efficiency, we opted for the Leave-One-Subject-Out (LOSO) methodology to assess the deep learning model’s average classification accuracy. Employing this approach, we conducted 20 iterations of training and testing for each input combination. During each iteration, one participant out of the 20 was reserved for testing, while the PLV features from the remaining 19 participants were utilized for model training. Following 50 epochs of model training, we identified the most effective model based on its classification accuracy on a validation set. This validation set, comprising 240 randomly selected vectors, equally distributed between remission and non-remission groups, was drawn from the training data. The model exhibiting the highest classification accuracy on the validation set underwent further testing.

## 3. Results

### 3.1 Clinical outcome

In the active tDCS treatment arm, 6 participants attained clinical remission (mean HAMD score: 2.83, SD 2.32) and 5 participants did not achieve clinical remission (mean HAMD score: 13.80, SD 3.83). In the sham treatment arm, 1 participant achieved clinical remission (HAMD score: 7) and 7 participants did not achieve clinical remission (mean HAMD score: 11.9, SD 2.54).

### 3.2 Effects of Treatment in EEG PLV connectivity

A significant main effect of treatment group was observed in full PLV between AF8-TP9 (*F* = 4.83, FDR-adjusted *p* = 0.03) and AF8-TP10 (*F* = 4.69, FDR-adjusted *p* = 0.04). Post-hoc tests revealed that the active treatment group showed increased PLV as compared to the sham treatment group at the end of treatment between AF8-TP9 (*t* = 2.31, FDR-adjusted *p* = 0.04) and AF8-TP10 (*t* = 2.68, FDR-adjusted *p* = 0.02), while there were no differences between groups at baseline between AF8-TP9 (*t* = 1.44, FDR-adjusted *p* = 0.17) and AF8-TP10 (*t* = 1.32, FDR-adjusted *p* = 0.21)(Figure 3, Supplementary Table 2).

In specific frequency bands, a significant main effect of group was found in beta band PLV between TP9-TP10 (*F* = 6.05, FDR-adjusted *p* = 0.02). Post-hoc tests revealed that at pre-treatment, the active treatment group showed a lower beta band PLV as compared to the sham group (*t* = -2.39, FDR-adjusted *p* = 0.03), while there was no significant difference at post-treatment (*t* = -1.11, FDR-adjusted *p* = 0.28).

Significant main effects of group were also observed in gamma band PLV between AF7-AF8 (*F* = 4.19, FDR-adjusted *p* = 0.05), AF7-TP9 (*F* = 6.80, FDR-adjusted *p* = 0.01), AF8-TP9 (*F* = 9.81, FDR-adjusted *p* = 0.00), AF8-TP10 (*F* = 7.78, FDR-adjusted *p* = 0.01) and TP9-TP10 (*F* = 5.12, FDR-adjusted *p* = 0.03) (Figure 3, Supplementary Table 1).

Post-hoc tests showed increased gamma PLV between AF7-AF8 in the active group as compared to the sham group which approached significance at post-treatment (*t* = 2.07, FDR-adjusted *p* = 0.06), while there were no significant differences at baseline (*t* = 1.32, FDR-adjusted *p* = 0.21). Similarly, post-hoc tests showed increased PLV post-treatment in the active group as compared to the sham group at AF8-TP9 (*t* = 3.03, FDR-adjusted *p* = 0.008) and AF8-TP10 (*t* = 3.00, FDR-adjusted *p* = 0.008), while there were no significant differences between groups at baseline at AF8-TP9 (*t* = 1.86, FDR-adjusted *p* = 0.08) and AF8-TP10 (*t* = 1.54, FDR-adjusted *p* = 0.14).

Post-hoc tests showed increased gamma PLV between AF7-TP9 at baseline in the active group (*t* = 2.40, FDR-adjusted *p* = 0.03) as compared to the sham group, while there were no significant differences between groups at post-treatment (*t* = 1.76, FDR-adjusted *p* = 0.10). Similarly, between TP9-TP10, post-hoc tests showed increased gamma PLV at baseline in the active group as compared to the sham groups which approached significance (*t* = 2.08, FDR-adjusted *p* = 0.06), but no differences between groups post-treatment (*t* = 0.69, FDR-adjusted *p* = 0.50).

There were no significant main effects of time or interaction effects (Supplementary Table 1).

### 3.3 Effects of Treatment in EEG power

There were no significant main effects of group or time or any interaction effects (Supplementary Table 1).

### 3.4 Relationship between changes in depression severity and EEG PLV connectivity

In the active treatment group, positive correlations with improvements in depressive symptoms following tDCS treatment were observed in full band PLV across several electrode pairs: AF7-AF8 (*R²* =0.73, FDR-adjusted p = 0.04), AF8-TP9 (*R²* = 0.74, FDR-adjusted *p* = 0.04), AF8-TP10 (*R²* = 0.82, FDR-adjusted *p* = 0.02), and TP9-TP10 (*R²* = 0.81, FDR-adjusted *p* = 0.01). Positive correlations were also observed in delta band PLV in the electrode pairs: AF8-TP10 (*R²* = 0.79, FDR-adjusted *p* = 0.02) and TP9-TP10 (*R²* = 0.72, FDR-adjusted *p* = 0.02). In the theta band PLV, positive correlations were found in the electrode pairs: AF7-AF8 (*R²* = 0.79, FDR-adjusted *p* = 0.02), AF7-TP9 (*R²* = 0.85, FDR-adjusted *p* = 0.01), AF7-TP10 (*R²* = 0.86, FDR-adjusted *p* = 0.01) and AF8-TP9 (*R²* = 0.88, FDR-adjusted *p* = 0.00). In alpha band PLV, a positive correlation was observed in the electrode pair AF8-TP9 (*R²* = 0.81, FDR-adjusted *p* = 0.01), and in beta band PLV, a positive correlation was found in electrode pair AF7-TP9 (*R²* = 0.88, FDR-adjusted *p* = 0.00). No regions showed a negative correlation with improvement in depressive symptoms. No significant correlations were found in the sham treatment group (Supplementary Table 4).

### 3.5 Relationship between changes in depression severity and EEG power

No significant correlation was found between change in EEG power and proportional change in HAMD scores from baseline to post-treatment across all participants (Supplementary Table 4).

### 3.6 Within group effects over time in active and sham groups

In the sham treatment group, significant reductions in alpha band PLV between AF7-AF8 (*t* = 2.51, FDR-adjusted p = 0.04), AF7-TP9 (t = 3.28, FDR-adjusted p = 0.01) and AF7-TP10 (t = 2.33, FDR-adjusted p = 0.05) and gamma PLV of AF7-AF8 (t = 3.17, FDR-adjusted p = 0.02) were found from baseline to post-treatment (Figure 1,Supplementary Table 3). In the active group, there were no significant differences from baseline to post-treatment.

**Figure 1.**
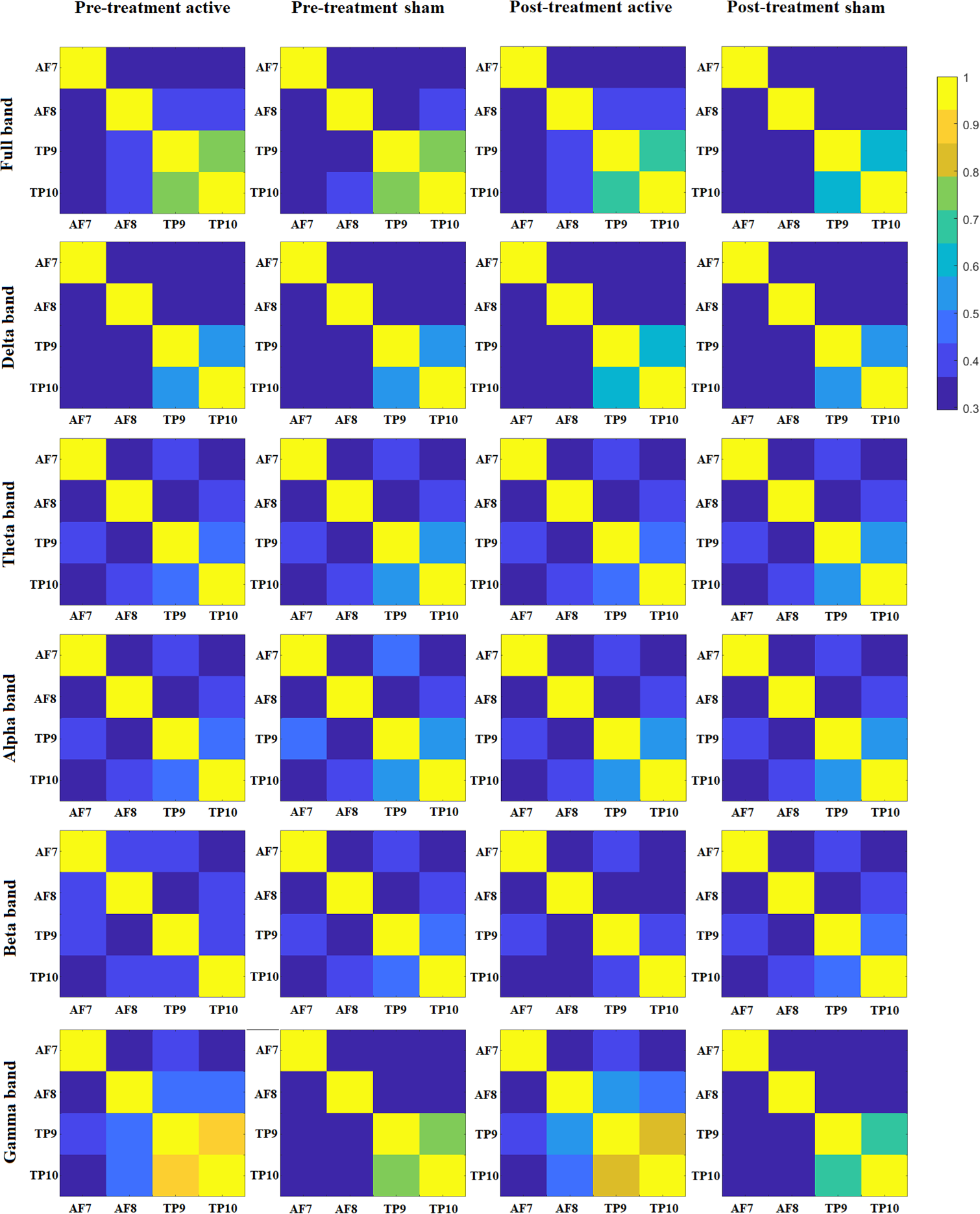
Image representing averaged PLV values across different group (active and sham group) and time (pre-and post-treatment). Rows depict EEG bands: full band, delta, theta, alpha, beta and gamma, and columns represent participant categories: pre-treatment active, pre-treatment sham, post-treatment active, and post-treatment sham groups.

### 3.7 Deep learning-based prediction

From EEG frequency bands, the prediction of treatment remission achieved the highest accuracies of: 60.04% (delta band), 67.46% (theta band), 68.42% (alpha band), 68.21% (beta band), 65.8% (gamma band), 46.48% (full band) with a 1DCNN model, except for the beta band in which the fully connected perceptron model achieved an accuracy of 71.19% (Supplementary Table 5).

Based on single EEG band PLV, prediction of treatment remission was the highest in alpha band PLV: 68.42% (sensitivity 14.29%, specificity of 100%). In all single band PLV classifications, specificity was significantly higher than sensitivity. PLV value matrix figure (Figure 2) shows connectivity patterns across EEG frequency bands.

**Figure 2.**
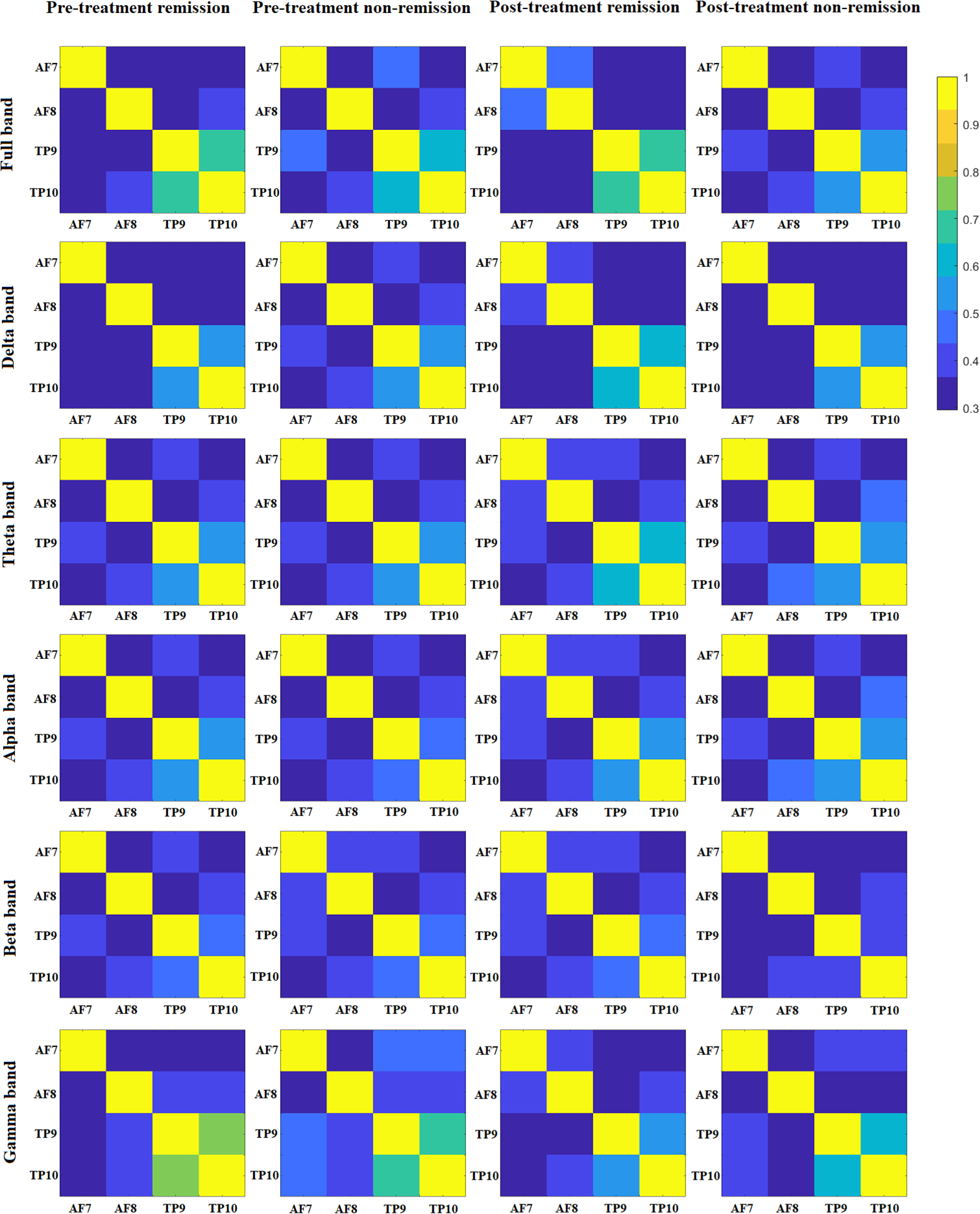
Image representing averaged PLV values across different group (remission and non-remission group) and time (pre- and post-treatment). Rows depict EEG bands: full band, delta, theta, alpha, beta and gamma, and columns represent participant categories: pre-treatment remission, pre-treatment non-remission, post-treatment remission, and post-treatment non-remission groups.

**Figure 3.**
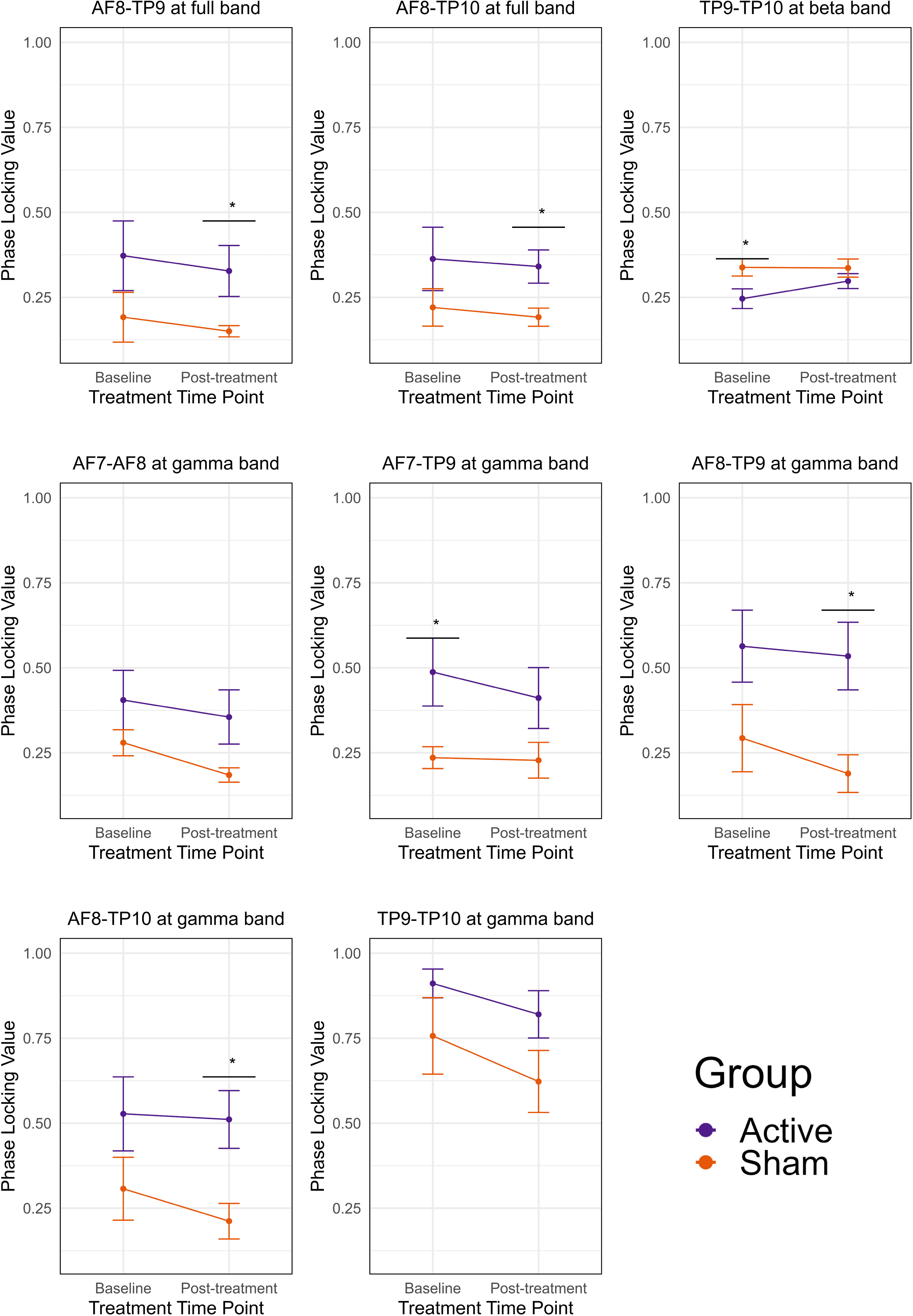
Boxplots presenting the PLV significant changes at different electrode pairs and frequency bands before and after treatment. The graphs show comparisons between the active treatment group (purple) and the sham treatment group (orange) at baseline and post-treatment time points. Significance between groups is denoted by *.

To enhance feature dimension and thereby provide more information for the deep learning models, we systematically combined the PLV features from multiple EEG bands. These high dimensional inputs were then utilized for predicting treatment outcomes. In combinations of two band PLV features, combination of theta and beta bands yielded the highest treatment remission prediction: 71.45% (sensitivity 53.29%, specificity 82.05%). In three EEG band PLV combinations, the highest treatment remission prediction accuracy was based on theta, alpha, and beta bands: 71.94% (sensitivity 52.88%, specificity of 83.06%). In four EEG band combinations, the combination of delta, theta, alpha, and beta bands yielded the highest treatment remission prediction accuracy: 66.32% (sensitivity 43.90%, specificity 79.40%). In the full five EEG bands delta, theta, alpha, beta, and gamma combination, treatment remission prediction accuracy was: 62.25% (sensitivity 54.48%, specificity 66.78%). The 1DCNN model consistently achieved the highest classification accuracy compared to the fully connected perceptron network in all the high performing band combinations, except the combination of the four EEG bands: delta, theta, alpha, beta, where the fully connected perceptron network achieved the highest classification accuracy of 66.32% (sensitivity 43.90%, specificity 79.40%) (Supplementary Table 6).

## 4. Discussion

We sought to examine the neurophysiology of MDD and effects of active and sham home-based tDCS treatment as measured by resting-state EEG. We applied deep learning to investigate whether baseline EEG measures could predict clinical remission from tDCS treatment. We observed a main effect of group in gamma PLV in frontal and temporal channel pairs in which the active tDCS treatment group showed higher connectivity as compared to the sham group. Post-hoc analyses revealed that this was evident at the end of treatment, while there were no differences between groups at baseline. Similarly, following rTMS treatment, increased gamma phase-based connectivity has been associated with clinical response, in which MDD participants who achieved a clinical response to rTMS treatment showed an increase in gamma phase-based connectivity, which was positively associated with improvements in mood and cognitive function (Bailey et al., 2018; Zuchowicz et al., 2019).

Gamma connectivity, reflecting synchronized brain activity across large-scale networks, is likely to be related to default mode network function (Buzsaki & Draguhn, 2004; Danilova, 2008; Fries et al., 2008; Arikan et al., 2019; Krukow & Jonak, 2022). Altered gamma connectivity has been observed in MDD, with hypoconnectivity within the frontoparietal network and hyperconnectivity within the default network, potentially reflect pathogenic mechanisms (Kaiser et al., 2015; Fitzgerald & Watson, 2018). In MDD, resting-state gamma oscillations showed a negative correlation with depressive symptoms, in particular with sleep and cognitive impairments (Liu et al., 2022). In contrast, increased gamma activity in the left prefrontal region in rTMS-responsive MDD patients has been linked to clinical improvements (Noda et al. 2017). In healthy individuals, enhanced gamma connectivity has been observed following mindfulness training, purported to reflect emotion regulation (Ng et al., 2023). The increased gamma connectivity observed in the active treatment as compared to the sham treatment group could reflect a mechanism of tDCS effects.

Within the active tDCS treatment group, we found significant positive correlations between changes in delta, theta, alpha, and beta PLV and improvement in depression severity. In support, clinical remission following rTMS treatment is associated increased delta, theta, alpha, and gamma PLV in multiple channel pairs (Zuchowicz et al., 2019), suggesting that these bands might distinguish the mechanisms between active and sham tDCS treatment.

To predict treatment remission, we applied deep learning networks to the PLV features extracted from the baseline EEG signals. Deep learning models are capable of handling high-dimensional data, such as EEG signals, which are often complex and non-linear. These models can automatically learn and extract useful features from raw EEG data without the need for manual feature extraction, which is a significant advantage over traditional machine learning methods. Scalogram images generated from EEGs have been used to train convolutional neural networks to classify MDD responders and non-responders to SSRI antidepressant treatment (Shahabi et al., 2021). In the present study, the highest prediction accuracy was achieved by combining PLV features from multiple EEG bands: theta, alpha, and beta. Notably, theta PLV exhibited the highest sensitivity, indicating that the classifier could predict treatment remission. Theta PLV also showed the strongest positive correlation between with an improvement in depressive symptoms. The significant individual-level associations between theta PLV changes and depression symptoms suggest that theta PLV could serve as a valuable predictor of tDCS treatment remission at an individual level.

Increased resting theta activity at baseline in the anterior cingulate cortex (ACC) is a potential predictor of treatment responses to antidepressant medication (Pizzagalli et al., 2018) as well as to rTMS treatment (Narushima et al., 2010). Theta activity reflects neural processes related to cognitive control and emotional regulation (Cavanagh et al., 2015), which are typically impaired in MDD (Grin-Yatsenko et al., 2010) and decreased resting-state theta connectivity has been observed MDD which was negatively correlated with depressive severity (Saletu et al., 2010; McVoy et al. 2019). Beta band power at baseline predicted rTMS treatment response in MDD, achieving 91% classification accuracy (Hasanzadeh et al., 2019), and baseline alpha power was predictive of treatment response to pharmacotherapy (Jaworska et al., 2019; Zhdanov et al., 2020).

As potential predictors of treatment response, power features have been utilised (Al-Kaysi et al., 2017; Hasanzadeh et al., 2019; Jaworska et al., 2019; Zhdanov et al., 2020), while connectivity features have been relatively underexamined. Alpha spectral correlation-based connectivity was used to predict rTMS treatment response, achieving 69.30% accuracy (Corlier et al., 2019), and directed transfer function (Korzeniewska et al., 2003) connectivity matrices of EEG bands has been applied to predict treatment remission to selective serotonin reuptake inhibitors (SSRIs) (Mirjebreili et al., 2024). In the present study, the predictive accuracy achieved from EEG band connectivity indicates its feasibility. Our findings complement previous studies of baseline resting-state EEG predicting treatment response to rTMS (Wozniak-Kwasniewska et al., 2015) and tDCS (Al-Kaysi et al., 2017). Together, the promising results suggest that baseline resting-state EEG may hold predictive value for treatment outcomes of non-invasive brain stimulation in MDD.

Limitations of the present study include the small sample size which limited investigation of neural markers that can distinguish response to active and placebo-sham treatment (Fu et al., 2024). The EEG data were acquired during a a resting state rather than during cognitive tasks, which prevents linking the activity to specific cognitive process. In the deep learning analysis, we applied leave-one-subject-out testing to mitigate overfitting, however, a more robust approach would be to test in a wholly independent sample. The specificity of the predictive models was often higher than the sensitivity, indicating that they were better at identifying participants who were more likely to have persistent depressive symptoms following treatment. This is consistent with EEG predictive models to rTMS and antidepressant treatments (Watts et al., 2022), suggesting a potential biomarker for non-response or treatment resistant depression and pointing towards the need for alternative or combination of treatments earlier in the course of treatment trials to prevent a series of treatment failures (Fu et al, 2024).

In summary, we examined the neurophysiological effects of a 10-week home-based tDCS treatment as measured by resting-state EEG-based functional connectivity and power measures. We found that the active tDCS group exhibited heightened gamma connectivity in frontal and temporal regions compared to the placebo-sham group, suggesting a neurophysiological mechanism of active tDCS that modulating neural network dynamics. Theta PLV demonstrated the strongest positive correlation with symptom severity improvement, suggesting a potential to predict individual-level treatment outcomes with tDCS. While integrating PLV features from various EEG frequency bands in the deep learning model generated the highest accuracy for predicting treatment remission, indicating that prediction response is distributed across multiple EEG bands.

## Data Availability

All data produced in the present study are available upon reasonable request to the authors

