## Supplementary Materials for "Home-based transcranial direct current stimulation (tDCS) in major depressive disorder: enhanced network synchronization with active relative to sham and deep learning-based predictors of remission"

**Treatment Group Differences in Demographic and Clinical Characteristics**

To assess the differences between the active and sham groups in variables such as age, gender, HAMD, and other potential covariates, we performed statistical tests, two sample t test across the groups for continuous variables (e.g., age and HAMD overall score), chi-square tests for categorical variables (e.g., sex).

The Welch Two Sample t-test was employed to assess the distinction in age between the Active and Sham groups. The findings indicated a lack of significance (t = 0.95347, p = 0.3555), suggesting no notable difference in age between the treatment groups. The mean age estimate for the Active group was 38.45, compared to 34.13 for the Sham group. Additionally, the Welch Two Sample t-test was utilized to evaluate disparities in baseline HAMD scores between the Active and Sham groups. The analysis yielded a non-significant outcome (t = 1.8132, p = 0.08752), indicating no statistically significant discrepancy in baseline HAMD scores between the treatment groups. The estimated mean baseline HAMD score for the Active group was 19.000, whereas for the Sham group, it was 17.625.

The chi-squared test with Yates' continuity correction was employed to examine the association between sex and treatment group in the dataset. The analysis revealed a non-significant result (χ² = 0.94573, df = 1, p = 0.3308), indicating that there is no substantial association between sex and the assignment to active or sham treatment groups. This suggests that the distribution of gender is similar across both treatment groups, and any observed differences are likely due to random chance rather than a systematic relationship. Baseline demographic and medication profiles did not differ between active treatment and placebo groups, suggesting that these factors are not predictive of treatment, and that demographic and medication effects did not confound the current results.

**Band power value calculation**

For the extraction of the EEG band power value $P$ from the windowed and band-passed EEG signals of length $N$, we employ Equation (1).

$P_{x}= \frac{1}{N}\left( x^{T}. x \right)$	(1)

In equation (1), $x$ represents the windowed and band-pass filtered EEG signal from an electrode, depicted as a column vector, while $x^{T}$ denotes its transpose. The power values are computed and subsequently averaged over all the windows of each participant for a specific frequency band, thereby yielding the final power value corresponding to that band. Consequently, there will be six power values per participant, each corresponding to a distinct frequency band.

**PLV calculation**

For PLV, the EEG signals are filtered into six EEG bands as above for the band power calculation. Within each band, the average PLV was calculated for the electrode pairs specified in the 6 pairwise electrodes comparisons.

The phase locking value (PLV) measures the synchrony between two time series signals (Hoke et al., 1989; Lachaux et al., 11999). It is one of the common measures used find functional connectivity between EEG signals from two electrodes. It calculates the long-range synchronization of neural activity from EEG data and express a measure of connectivity. The PLV is a statistical quantity, bounded between 0 and 1. The PLV value near 1 indicates phase difference varies little across the two EEG signals and a value near 0 indicates otherwise. The single trial $PLV$ over $N$ samples is calculated as follows,

$PLV = \left| \frac{1}{N}\Sigma_{k=1}^{N}e^{i\Delta\theta\left( k \right)} \right|$	(2)

$\Delta\theta\left( k \right) = \theta_{1}\left( k \right)-\theta_{2}\left( k \right)$	(3)

In the above equations, $\theta_{1}\left( k \right)$ and $\theta_{2}\left( k \right)$ are instantaneous phase of two analytical signals derived from their corresponding EEG signals. The analytical signals and are computed from the real signals and using the Hilbert transform (Boashash, 2003). There were 72 (6 comparisons X 6 frequency bands X 2 Treatment Time Point) dependent measures for PLV.

**Quantitative Statistical Analysis**

To determine whether tDCS treatment influences the overall severity of depression, we performed a two-way ANOVA on the total HAMD score. The HAMD score was considered the dependent variable, while the treatment group (active or sham) and the treatment time point (baseline or post-treatment) were as the independent variables.

To assess whether variations in EEG measures during treatment were associated with alterations in depression severity, we performed a Peason’s correlation analysis between change in EEG measures after 10-week treatment and proportional change in HAMD from baseline to 10-week treatment across subjects. The proportional change in HAMD is determined by calculating the absolute difference between the HAMD score after 10 weeks of treatment and the baseline HAMD score. This absolute difference is then divided by the baseline HAMD score, providing a measure of relative change in depression severity over the treatment period.

To test whether EEG measures in general might change in response to tDCS, we performed a two-way fixed-effects ANOVA for each EEG variable, with EEG variable as the dependent variable and Treatment Group (active/sham) and Time Point (pre- or post-treatment) as independent variables. We conducted a total of 60 statistical tests.

Dependent measure ~ Treatment Group * Time Point

The coefficients were estimated in the R statistical environment using linear regression (*lm* built-in function). For each test, *η^2^* associated with each independent variable (treatment group and time point) was calculated to measure effect size.

**Time-Dependent Dynamics: Two-Way ANOVA Reveals Time Effect on HAMD Overall Score**

The statistical analysis revealed a significant main effect of Time Point (F = 51.79, p < 0.001), indicating a substantial overall impact of time on the dependent variable (i.e., HAMD overall score). However, the main effect of Group did not reach significance (F = 0.64, p = 0.43). The interaction effect between Group and Time Point approached significance (F = 3.48, p = 0.07).

The paired two-sample t-tests were employed to examine alterations in HAMD overall scores within each treatment group across the time point (i.e., baseline and post-treatment). In the Active Group, a substantial reduction in HAMD scores was identified, with a mean difference of 11.18182 (t = 5.896, df = 10, p = 0.0001519, 95% CI: 6.956163 to 15.407473). Likewise, Group 2 exhibited a noteworthy decrease in HAMD scores, with a mean difference of 6.375 (t = 4.7736, df = 7, p = 0.002027, 95% CI: 3.217113 to 9.532887). The adjusted p-values, corrected for False Discovery Rate, were applied to mitigate the risk of Type I errors in multiple comparisons.

Supplementary Materials

**Supplementary Table 1.**

Means, Standard Deviations, and main effect of Treatment Group (active/sham), main effect of time and interaction from Two-Way Analyses of Variance in Power and Phase Locking Value (PLV)

| Variable | Main effect of Group | | | | | | Main effect of Time | | | | | | Interaction between group and time | |
| --- | --- | --- | --- | --- | --- | --- | --- | --- | --- | --- | --- | --- | --- | --- |
|  | Active | | Sham | | *F (*1,1) | *p* | Baseline | | Post-treatment | | *F (*1,1) | *p* | *F (*1,1) | *p* |
|  | *M* | *SD* | *M* | *SD* |  |  | *M* | *SD* | *M* | *SD* |  |  |  |  |
| Full Power |  |  |  |  |  |  |  |  |  |  |  |  |  |  |
| AF7 | 3251.92 | 9264.95 | 3832.00 | 10241.05 | 0.32 | 0.81 | 2620.99 | 7657.80 | 2396.58 | 7432.07 | 0.01 | 0.93 | 1.21 | 0.32 |
| AF8 | 4646.51 | 13784.14 | 1410.51 | 2160.54 | 0.63 | 0.60 | 794.74 | 1469.15 | 3318.58 | 10960.87 | 0.92 | 0.34 | 0.57 | 0.64 |
| TP9 | 9554.02 | 13780.02 | 12516.48 | 27757.36 | 0.35 | 0.79 | 4052.38 | 8383.25 | 13074.02 | 24189.30 | 2.05 | 0.16 | 0.07 | 0.98 |
| TP10 | 12261.22 | 31192.75 | 9667.66 | 26569.95 | 0.17 | 0.91 | 10517.78 | 25953.12 | 7175.61 | 24402.48 | 0.15 | 0.70 | 0.65 | 0.59 |
| Delta Power |  |  |  |  |  |  |  |  |  |  |  |  |  |  |
| AF7 | 358.23 | 586.55 | 327.06 | 595.73 | 0.03 | 0.87 | 279.34 | 476.81 | 410.88 | 679.08 | 0.47 | 0.50 | 1.53 | 0.22 |
| AF8 | 329.83 | 514.35 | 216.67 | 383.17 | 0.53 | 0.47 | 256.36 | 480.75 | 308.00 | 452.84 | 0.11 | 0.74 | 0.30 | 0.59 |
| TP9 | 333.19 | 328.22 | 465.31 | 820.20 | 0.47 | 0.50 | 271.79 | 374.65 | 505.85 | 725.26 | 1.50 | 0.23 | 0.16 | 0.69 |
| TP10 | 399.02 | 589.75 | 425.08 | 957.27 | 0.01 | 0.92 | 408.42 | 876.64 | 411.56 | 633.44 | 0.00 | 0.99 | 1.81 | 0.19 |
| Theta Power |  |  |  |  |  |  |  |  |  |  |  |  |  |  |
| AF7 | 90.58 | 125.42 | 71.32 | 118.92 | 0.22 | 0.64 | 71.18 | 108.66 | 93.76 | 135.10 | 0.31 | 0.58 | 0.56 | 0.46 |
| AF8 | 63.68 | 70.60 | 65.50 | 123.80 | 0.00 | 0.95 | 44.72 | 58.39 | 84.17 | 119.83 | 1.66 | 0.21 | 1.85 | 0.18 |
| TP9 | 82.63 | 71.06 | 171.52 | 319.05 | 1.53 | 0.22 | 105.96 | 242.50 | 134.15 | 188.57 | 0.16 | 0.69 | 0.03 | 0.87 |
| TP10 | 96.65 | 141.36 | 96.11 | 164.27 | 0.00 | 0.99 | 82.06 | 138.23 | 110.79 | 162.03 | 0.33 | 0.57 | 0.62 | 0.44 |
| Alpha Power |  |  |  |  |  |  |  |  |  |  |  |  |  |  |
| AF7 | 55.23 | 122.31 | 58.65 | 127.29 | 0.01 | 0.94 | 73.68 | 167.82 | 39.66 | 46.78 | 0.69 | 0.41 | 0.02 | 0.90 |
| AF8 | 46.07 | 68.65 | 38.16 | 65.59 | 0.13 | 0.72 | 28.68 | 39.55 | 56.80 | 84.46 | 1.68 | 0.20 | 0.94 | 0.34 |
| TP9 | 40.26 | 29.83 | 91.27 | 140.82 | 2.60 | 0.12 | 57.22 | 107.90 | 66.25 | 84.95 | 0.08 | 0.77 | 0.00 | 0.98 |
| TP10 | 48.11 | 52.83 | 61.23 | 81.50 | 0.35 | 0.56 | 42.47 | 44.98 | 64.81 | 81.20 | 1.05 | 0.31 | 0.02 | 0.88 |
| Beta Power |  |  |  |  |  |  |  |  |  |  |  |  |  |  |
| AF7 | 300.18 | 900.79 | 384.48 | 1268.19 | 0.06 | 0.81 | 537.73 | 1471.21 | 133.62 | 199.80 | 1.34 | 0.26 | 0.20 | 0.66 |
| AF8 | 228.00 | 501.95 | 85.73 | 102.65 | 1.20 | 0.28 | 112.46 | 209.59 | 223.73 | 512.53 | 0.75 | 0.39 | 0.01 | 0.91 |
| TP9 | 104.57 | 110.27 | 144.35 | 241.78 | 0.44 | 0.51 | 120.83 | 223.86 | 121.80 | 116.64 | 0.00 | 0.99 | 0.18 | 0.68 |
| TP10 | 107.37 | 121.14 | 86.68 | 86.35 | 0.33 | 0.57 | 82.54 | 102.82 | 114.78 | 111.52 | 0.82 | 0.37 | 0.00 | 0.95 |
| Gamma Power |  |  |  |  |  |  |  |  |  |  |  |  |  |  |
| AF7 | 3010.82 | 9379.19 | 293.08 | 808.83 | 1.25 | 0.27 | 1845.12 | 6969.21 | 1887.90 | 7642.39 | 0.00 | 0.99 | 0.02 | 0.89 |
| AF8 | 2877.07 | 10837.26 | 113.23 | 113.80 | 1.02 | 0.32 | 428.01 | 1262.05 | 2998.70 | 11656.83 | 0.90 | 0.35 | 0.61 | 0.44 |
| TP9 | 11912.06 | 23548.25 | 5075.84 | 15660.21 | 1.03 | 0.32 | 3976.27 | 9446.09 | 14091.03 | 27057.98 | 2.30 | 0.14 | 0.00 | 0.94 |
| TP10 | 11984.07 | 32326.42 | 5853.44 | 20910.80 | 0.42 | 0.52 | 11388.49 | 28798.31 | 7417.01 | 27662.66 | 0.18 | 0.67 | 0.28 | 0.60 |
| Full PLV |  |  |  |  |  |  |  |  |  |  |  |  |  |  |
| AF7-AF8 | 0.24 | 0.19 | 0.19 | 0.09 | 0.88 | 0.35 | 0.22 | 0.18 | 0.22 | 0.13 | 0.00 | 0.96 | 0.90 | 0.35 |
| AF7-TP9 | 0.29 | 0.23 | 0.18 | 0.06 | 3.06 | 0.09 | 0.24 | 0.17 | 0.24 | 0.21 | 0.01 | 0.94 | 0.09 | 0.77 |
| AF7-TP10 | 0.22 | 0.20 | 0.13 | 0.05 | 2.54 | 0.12 | 0.18 | 0.18 | 0.18 | 0.14 | 0.00 | 0.98 | 0.00 | 0.96 |
| **AF8-TP9** | **0.35** | **0.29** | **0.17** | **0.15** | **4.83** | **0.03** | 0.25 | 0.21 | 0.30 | 0.30 | 0.29 | 0.59 | 0.00 | 0.98 |
| **AF8-TP10** | **0.35** | **0.24** | **0.21** | **0.12** | **4.69** | **0.04** | 0.28 | 0.15 | 0.30 | 0.26 | 0.14 | 0.71 | 0.00 | 0.96 |
| TP9-TP10 | 0.69 | 0.28 | 0.61 | 0.23 | 0.73 | 0.40 | 0.60 | 0.25 | 0.71 | 0.27 | 1.43 | 0.24 | 0.01 | 0.93 |
| Delta PLV |  |  |  |  |  |  |  |  |  |  |  |  |  |  |
| AF7-AF8 | 0.31 | 0.09 | 0.33 | 0.12 | 0.36 | 0.55 | 0.31 | 0.10 | 0.33 | 0.11 | 0.39 | 0.54 | 0.66 | 0.42 |
| AF7-TP9 | 0.32 | 0.08 | 0.33 | 0.09 | 0.40 | 0.53 | 0.32 | 0.08 | 0.33 | 0.08 | 0.01 | 0.91 | 0.20 | 0.66 |
| AF7-TP10 | 0.27 | 0.08 | 0.31 | 0.11 | 1.64 | 0.21 | 0.29 | 0.09 | 0.29 | 0.10 | 0.01 | 0.93 | 0.07 | 0.79 |
| AF8-TP9 | 0.32 | 0.12 | 0.32 | 0.11 | 0.02 | 0.88 | 0.32 | 0.11 | 0.32 | 0.13 | 0.03 | 0.87 | 0.62 | 0.44 |
| AF8-TP10 | 0.32 | 0.10 | 0.34 | 0.09 | 0.39 | 0.54 | 0.33 | 0.08 | 0.33 | 0.11 | 0.00 | 0.99 | 0.52 | 0.47 |
| TP9-TP10 | 0.55 | 0.14 | 0.58 | 0.15 | 0.19 | 0.67 | 0.58 | 0.14 | 0.55 | 0.15 | 0.42 | 0.52 | 0.30 | 0.59 |
| Theta PLV |  |  |  |  |  |  |  |  |  |  |  |  |  |  |
| AF7-AF8 | 0.25 | 0.14 | 0.23 | 0.11 | 0.25 | 0.62 | 0.22 | 0.11 | 0.27 | 0.14 | 0.93 | 0.34 | 0.60 | 0.44 |
| AF7-TP9 | 0.33 | 0.09 | 0.32 | 0.11 | 0.01 | 0.92 | 0.32 | 0.10 | 0.33 | 0.10 | 0.13 | 0.72 | 1.10 | 0.30 |
| AF7-TP10 | 0.23 | 0.11 | 0.23 | 0.09 | 0.00 | 0.97 | 0.22 | 0.07 | 0.25 | 0.12 | 0.75 | 0.39 | 0.38 | 0.54 |
| AF8-TP9 | 0.24 | 0.14 | 0.23 | 0.10 | 0.09 | 0.76 | 0.23 | 0.11 | 0.25 | 0.13 | 0.30 | 0.59 | 0.02 | 0.89 |
| AF8-TP10 | 0.33 | 0.11 | 0.32 | 0.11 | 0.08 | 0.78 | 0.32 | 0.10 | 0.33 | 0.13 | 0.00 | 0.96 | 0.23 | 0.63 |
| TP9-TP10 | 0.41 | 0.16 | 0.48 | 0.14 | 1.90 | 0.18 | 0.44 | 0.16 | 0.45 | 0.16 | 0.12 | 0.73 | 0.85 | 0.36 |
| Alpha PLV |  |  |  |  |  |  |  |  |  |  |  |  |  |  |
| AF7-AF8 | 0.28 | 0.18 | 0.21 | 0.10 | 2.11 | 0.16 | 0.23 | 0.16 | 0.27 | 0.15 | 0.72 | 0.40 | 1.23 | 0.28 |
| AF7-TP9 | 0.36 | 0.11 | 0.35 | 0.11 | 0.12 | 0.73 | 0.34 | 0.09 | 0.37 | 0.13 | 0.88 | 0.36 | 2.18 | 0.15 |
| AF7-TP10 | 0.23 | 0.12 | 0.20 | 0.08 | 0.67 | 0.42 | 0.20 | 0.10 | 0.24 | 0.11 | 0.97 | 0.33 | 1.10 | 0.30 |
| AF8-TP9 | 0.24 | 0.12 | 0.21 | 0.09 | 0.58 | 0.45 | 0.22 | 0.11 | 0.24 | 0.11 | 0.22 | 0.64 | 0.75 | 0.39 |
| AF8-TP10 | 0.35 | 0.10 | 0.33 | 0.12 | 0.29 | 0.59 | 0.33 | 0.10 | 0.35 | 0.12 | 0.10 | 0.75 | 0.44 | 0.51 |
| TP9-TP10 | 0.44 | 0.16 | 0.49 | 0.13 | 1.15 | 0.29 | 0.46 | 0.15 | 0.47 | 0.15 | 0.02 | 0.89 | 2.57 | 0.12 |
| Beta PLV |  |  |  |  |  |  |  |  |  |  |  |  |  |  |
| AF7-AF8 | 0.23 | 0.16 | 0.17 | 0.06 | 2.02 | 0.16 | 0.19 | 0.12 | 0.23 | 0.14 | 0.76 | 0.39 | 0.31 | 0.58 |
| AF7-TP9 | 0.25 | 0.08 | 0.25 | 0.08 | 0.00 | 0.95 | 0.24 | 0.06 | 0.26 | 0.09 | 0.46 | 0.50 | 0.09 | 0.76 |
| AF7-TP10 | 0.16 | 0.07 | 0.15 | 0.08 | 0.15 | 0.70 | 0.16 | 0.09 | 0.16 | 0.07 | 0.05 | 0.83 | 0.00 | 0.96 |
| AF8-TP9 | 0.15 | 0.07 | 0.14 | 0.06 | 0.50 | 0.48 | 0.14 | 0.06 | 0.15 | 0.07 | 0.19 | 0.66 | 0.13 | 0.72 |
| AF8-TP10 | 0.24 | 0.08 | 0.23 | 0.08 | 0.12 | 0.73 | 0.22 | 0.06 | 0.24 | 0.10 | 0.45 | 0.51 | 0.00 | 0.99 |
| **TP9-TP10** | **0.27** | **0.09** | **0.34** | **0.07** | **6.05** | **0.02** | 0.31 | 0.07 | 0.28 | 0.10 | 1.25 | 0.27 | 1.03 | 0.32 |
| Gamma PLV |  |  |  |  |  |  |  |  |  |  |  |  |  |  |
| **AF7-AF8** | **0.38** | **0.27** | **0.23** | **0.10** | **4.19** | **0.05** | 0.28 | 0.22 | 0.35 | 0.24 | 0.93 | 0.34 | 0.10 | 0.76 |
| **AF7-TP9** | **0.45** | **0.31** | **0.23** | **0.12** | **6.80** | **0.01** | 0.33 | 0.26 | 0.38 | 0.28 | 0.33 | 0.57 | 0.17 | 0.68 |
| AF7-TP10 | 0.39 | 0.29 | 0.25 | 0.16 | 3.12 | 0.09 | 0.30 | 0.24 | 0.36 | 0.26 | 0.54 | 0.47 | 0.71 | 0.41 |
| **AF8-TP9** | **0.55** | **0.33** | **0.24** | **0.23** | **9.81** | **0.00** | 0.39 | 0.32 | 0.45 | 0.34 | 0.39 | 0.54 | 0.15 | 0.70 |
| **AF8-TP10** | **0.52** | **0.32** | **0.26** | **0.21** | **7.78** | **0.01** | 0.39 | 0.27 | 0.43 | 0.33 | 0.29 | 0.59 | 0.18 | 0.67 |
| **TP9-TP10** | **0.87** | **0.19** | **0.69** | **0.29** | **5.12** | **0.03** | 0.74 | 0.26 | 0.85 | 0.24 | 2.02 | 0.16 | 0.08 | 0.78 |

*Note. ^*^p < .05, ^**^p < .01, p values are corrected for False Discovery Rate (FDR)*

**Supplementary Materials**

**Table 2.**

*Results of Post-hoc test Examining the Treatment group difference at the Baseline and Post-treatment for variables with significant main effect of group from the previous Two-way Anova*

| PLV Variables | Baseline | | | |  |  | Post-Treatment | | | |  |  |
| --- | --- | --- | --- | --- | --- | --- | --- | --- | --- | --- | --- | --- |
|  | Active | | Sham | | *t* | *p* | Active | | Sham | | *t* | *p* |
|  | *M* | *SD* | *M* | *SD* |  |  | *M* | *SD* | *M* | *SD* |  |  |
| Full |  |  |  |  |  |  |  |  |  |  |  |  |
| AF8-TP9 | 0.37 | 0.34 | 0.19 | 0.21 | 1.44 | 0.17 | 0.33 | 0.25 | 0.15 | 0.05 | 2.31 | 0.04 |
| AF8-TP10 | 0.36 | 0.31 | 0.22 | 0.16 | 1.32 | 0.21 | 0.34 | 1.62 | 0.19 | 0.08 | 2.68 | 0.02 |
| Beta |  |  |  |  |  |  |  |  |  |  |  |  |
| TP9-TP10 | 0.25 | 0.10 | 0.34 | 0.07 | -2.39 | 0.03 | 0.30 | 0.07 | 0.34 | 0.08 | -1.11 | 0.28 |
| Gamma |  |  |  |  |  |  |  |  |  |  |  |  |
| AF7-AF8 | 0.41 | 0.29 | 0.28 | 0.11 | 1.32 | 0.21 | 0.36 | 0.26 | 0.18 | 0.06 | 2.07 | 0.06 |
| AF7-TP9 | 0.49 | 0.33 | 0.24 | 0.09 | 2.40 | 0.03 | 0.41 | 0.30 | 0.23 | 0.15 | 1.76 | 0.10 |
| AF8-TP9 | 0.56 | 0.35 | 0.29 | 0.28 | 1.86 | 0.08 | 0.53 | 0.33 | 0.19 | 0.16 | 3.03 | 0.008 |
| AF8-TP10 | 0.53 | 0.36 | 0.31 | 0.26 | 1.54 | 0.14 | 0.51 | 0.28 | 0.21 | 0.15 | 3.00 | 0.008 |
| TP9-TP10 | 0.45 | 0.31 | 0.24 | 0.10 | 2.08 | 0.06 | 0.33 | 0.26 | 0.26 | 0.21 | 0.69 | 0.50 |

*Note.* ^*^*p* < .05, ^**^*p* < .01, *p values are adjusted by FDR*

**Supplementary Materials**

**Supplementary Table 3.**

Change in PLV from the pre- to post-treatment within each treatment group (active/sham) from paired two-sample t test

| Variable | Active Group | | | | | | Sham Group | | | | | |
| --- | --- | --- | --- | --- | --- | --- | --- | --- | --- | --- | --- | --- |
|  | Baseline | | Post-Treatment | | *t* | *p* | Baseline | | Post-Treatment | | *t* | *p* |
|  | *M* | *SD* | *M* | *SD* |  |  | *M* | *SD* | *M* | *SD* |  |  |
| Full  Power |  |  |  |  |  |  |  |  |  |  |  |  |
| AF7 | 3722.30 | 9714.62 | 3308.89 | 9722.66 | 0.09 | 0.93 | 573.73 | 809.57 | 1675.11 | 3739.57 | -0.81 | 0.44 |
| AF8 | 5171.95 | 14376.98 | 1179.26 | 1857.49 | 0.93 | 0.37 | 770.21 | 911.58 | 266.02 | 263.37 | 1.40 | 0.21 |
| TP9 | 15580.99 | 27332.50 | 6220.20 | 10574.98 | 0.98 | 0.35 | 9626.95 | 20344.73 | 1071.63 | 1849.47 | 1.31 | 0.23 |
| TP10 | 11398.95 | 31993.68 | 10765.71 | 26182.16 | 0.06 | 0.95 | 1368.53 | 1545.61 | 10176.89 | 27431.72 | -0.92 | 0.39 |
| Delta Power |  |  |  |  |  |  |  |  |  |  |  |  |
| AF7 | 525.18 | 778.56 | 191.29 | 234.56 | 1.31 | 0.22 | 253.71 | 520.68 | 400.42 | 690.71 | -0.60 | 0.57 |
| AF8 | 319.77 | 438.44 | 339.88 | 602.59 | -0.15 | 0.89 | 291.82 | 502.18 | 141.52 | 222.54 | 0.71 | 0.50 |
| TP9 | 417.21 | 401.63 | 249.18 | 222.23 | 1.11 | 0.29 | 627.74 | 1045.37 | 302.88 | 537.09 | 1.63 | 0.15 |
| TP10 | 543.40 | 756.59 | 254.63 | 334.81 | 1.37 | 0.20 | 230.28 | 386.21 | 619.88 | 1314.44 | -0.82 | 0.44 |
| Theta Power |  |  |  |  |  |  |  |  |  |  |  |  |
| AF7 | 114.84 | 144.43 | 66.32 | 104.29 | 0.80 | 0.44 | 64.78 | 124.39 | 77.86 | 121.39 | -0.25 | 0.81 |
| AF8 | 65.62 | 72.71 | 61.74 | 71.92 | 0.16 | 0.88 | 109.68 | 167.59 | 21.32 | 17.25 | 1.44 | 0.19 |
| TP9 | 101.75 | 82.41 | 63.51 | 54.84 | 1.16 | 0.27 | 178.71 | 278.95 | 164.34 | 374.43 | 0.24 | 0.81 |
| TP10 | 127.69 | 183.20 | 65.62 | 79.27 | 1.30 | 0.22 | 87.55 | 136.02 | 104.66 | 197.87 | -0.21 | 0.84 |
| Alpha Power |  |  |  |  |  |  |  |  |  |  |  |  |
| AF7 | 40.45 | 38.40 | 70.02 | 171.64 | -0.57 | 0.58 | 38.56 | 59.32 | 78.73 | 174.02 | -0.62 | 0.56 |
| AF8 | 51.20 | 86.37 | 40.95 | 48.78 | 0.38 | 0.71 | 64.51 | 87.00 | 11.80 | 7.83 | 1.72 | 0.13 |
| TP9 | 44.41 | 32.18 | 36.10 | 28.20 | 0.69 | 0.50 | 96.28 | 123.69 | 86.27 | 164.73 | 0.37 | 0.72 |
| TP10 | 57.87 | 65.56 | 38.36 | 36.79 | 1.12 | 0.29 | 74.34 | 103.13 | 48.12 | 56.63 | 0.64 | 0.54 |
| Beta Power |  |  |  |  |  |  |  |  |  |  |  |  |
| AF7 | 164.30 | 250.74 | 436.06 | 1265.10 | -0.72 | 0.49 | 91.43 | 96.55 | 677.54 | 1800.20 | -0.92 | 0.39 |
| AF8 | 290.21 | 670.41 | 165.79 | 266.74 | 0.57 | 0.58 | 132.32 | 130.02 | 39.14 | 26.64 | 2.08 | 0.08 |
| TP9 | 115.64 | 110.78 | 93.50 | 113.99 | 0.51 | 0.62 | 130.27 | 131.56 | 158.42 | 327.88 | -0.33 | 0.75 |
| TP10 | 122.51 | 126.12 | 92.24 | 120.04 | 0.76 | 0.47 | 104.14 | 95.03 | 69.21 | 79.06 | 1.28 | 0.24 |
| Gamma Power |  |  |  |  |  |  |  |  |  |  |  |  |
| AF7 | 3168.88 | 10041.41 | 2852.76 | 9156.99 | 0.07 | 0.94 | 126.55 | 143.07 | 459.60 | 1148.05 | -0.82 | 0.44 |
| AF8 | 5064.20 | 15278.33 | 689.95 | 1638.81 | 0.94 | 0.37 | 158.63 | 141.86 | 67.83 | 54.01 | 1.89 | 0.10 |
| TP9 | 17169.86 | 31015.03 | 6654.25 | 11905.56 | 0.98 | 0.35 | 9857.63 | 21750.04 | 294.05 | 436.76 | 1.26 | 0.25 |
| TP10 | 12093.82 | 36313.35 | 11874.32 | 29594.06 | 0.02 | 0.99 | 986.41 | 1511.57 | 10720.48 | 29674.23 | -0.94 | 0.38 |
| Full PLV |  |  |  |  |  |  |  |  |  |  |  |  |
| AF7-AF8 | 0.22 | 0.15 | 0.26 | 0.23 | -0.58 | 0.57 | 0.22 | 0.11 | 0.16 | 0.06 | 2.18 | 0.07 |
| AF7-TP9 | 0.30 | 0.26 | 0.28 | 0.21 | 0.19 | 0.85 | 0.17 | 0.08 | 0.19 | 0.05 | -0.86 | 0.42 |
| AF7-TP10 | 0.22 | 0.18 | 0.22 | 0.23 | 0.04 | 0.97 | 0.13 | 0.03 | 0.13 | 0.07 | -0.06 | 0.95 |
| AF8-TP9 | 0.37 | 0.34 | 0.33 | 0.25 | 0.51 | 0.62 | 0.19 | 0.21 | 0.15 | 0.05 | 0.57 | 0.58 |
| AF8-TP10 | 0.36 | 0.31 | 0.34 | 0.16 | 0.22 | 0.83 | 0.22 | 0.16 | 0.19 | 0.08 | 0.49 | 0.64 |
| TP9-TP10 | 0.73 | 0.28 | 0.64 | 0.28 | 1.26 | 0.24 | 0.67 | 0.27 | 0.56 | 0.19 | 1.38 | 0.21 |
| Delta PLV |  |  |  |  |  |  |  |  |  |  |  |  |
| AF7-AF8 | 0.31 | 0.09 | 0.32 | 0.10 | -0.06 | 0.95 | 0.36 | 0.13 | 0.31 | 0.11 | 1.47 | 0.19 |
| AF7-TP9 | 0.31 | 0.07 | 0.32 | 0.09 | -0.23 | 0.83 | 0.34 | 0.10 | 0.32 | 0.08 | 0.58 | 0.58 |
| AF7-TP10 | 0.28 | 0.09 | 0.27 | 0.07 | 0.29 | 0.78 | 0.31 | 0.11 | 0.32 | 0.11 | -0.19 | 0.85 |
| AF8-TP9 | 0.33 | 0.15 | 0.30 | 0.10 | 0.64 | 0.53 | 0.31 | 0.11 | 0.34 | 0.12 | -0.84 | 0.43 |
| AF8-TP10 | 0.33 | 0.12 | 0.32 | 0.08 | 0.37 | 0.72 | 0.33 | 0.09 | 0.36 | 0.08 | -1.25 | 0.25 |
| TP9-TP10 | 0.53 | 0.16 | 0.58 | 0.12 | -1.22 | 0.25 | 0.58 | 0.14 | 0.58 | 0.17 | -0.01 | 0.99 |
| Theta PLV |  |  |  |  |  |  |  |  |  |  |  |  |
| AF7-AF8 | 0.26 | 0.14 | 0.25 | 0.15 | 0.23 | 0.82 | 0.27 | 0.15 | 0.19 | 0.04 | 1.69 | 0.14 |
| AF7-TP9 | 0.32 | 0.10 | 0.33 | 0.09 | -0.48 | 0.64 | 0.35 | 0.11 | 0.30 | 0.11 | 1.90 | 0.10 |
| AF7-TP10 | 0.24 | 0.13 | 0.23 | 0.08 | 0.25 | 0.81 | 0.26 | 0.11 | 0.21 | 0.05 | 1.53 | 0.17 |
| AF8-TP9 | 0.25 | 0.14 | 0.23 | 0.14 | 0.30 | 0.77 | 0.24 | 0.13 | 0.21 | 0.05 | 0.82 | 0.44 |
| AF8-TP10 | 0.32 | 0.13 | 0.34 | 0.10 | -0.26 | 0.80 | 0.33 | 0.14 | 0.31 | 0.10 | 0.59 | 0.58 |
| TP9-TP10 | 0.40 | 0.16 | 0.43 | 0.17 | -0.60 | 0.56 | 0.52 | 0.13 | 0.45 | 0.15 | 1.66 | 0.14 |
| Alpha PLV |  |  |  |  |  |  |  |  |  |  |  |  |
| AF7-AF8 | 0.28 | 0.18 | 0.28 | 0.19 | -0.08 | 0.94 | **0.26** | **0.12** | **0.15** | **0.02** | **2.51** | **0.04** |
| AF7-TP9 | 0.35 | 0.13 | 0.37 | 0.10 | -0.26 | 0.80 | **0.40** | **0.13** | **0.30** | **0.08** | **3.28** | **0.01** |
| AF7-TP10 | 0.23 | 0.13 | 0.23 | 0.12 | 0.07 | 0.95 | **0.24** | **0.10** | **0.16** | **0.01** | **2.33** | **0.05** |
| AF8-TP9 | 0.23 | 0.11 | 0.24 | 0.14 | -0.21 | 0.83 | 0.24 | 0.11 | 0.19 | 0.04 | 1.48 | 0.18 |
| AF8-TP10 | 0.34 | 0.11 | 0.35 | 0.10 | -0.22 | 0.83 | 0.35 | 0.14 | 0.31 | 0.09 | 1.09 | 0.31 |
| TP9-TP10 | 0.41 | 0.16 | 0.47 | 0.16 | -1.71 | 0.12 | 0.54 | 0.10 | 0.44 | 0.15 | 1.64 | 0.14 |
| Beta PLV |  |  |  |  |  |  |  |  |  |  |  |  |
| AF7-AF8 | 0.24 | 0.17 | 0.23 | 0.15 | 0.45 | 0.66 | 0.21 | 0.07 | 0.14 | 0.03 | 2.34 | 0.05 |
| AF7-TP9 | 0.26 | 0.10 | 0.25 | 0.07 | 0.30 | 0.77 | 0.27 | 0.09 | 0.24 | 0.07 | 1.34 | 0.22 |
| AF7-TP10 | 0.16 | 0.07 | 0.17 | 0.08 | -0.18 | 0.86 | 0.15 | 0.07 | 0.16 | 0.10 | -0.11 | 0.92 |
| AF8-TP9 | 0.16 | 0.08 | 0.15 | 0.07 | 0.13 | 0.90 | 0.15 | 0.08 | 0.13 | 0.05 | 0.83 | 0.44 |
| AF8-TP10 | 0.25 | 0.11 | 0.23 | 0.06 | 0.41 | 0.69 | 0.24 | 0.09 | 0.22 | 0.07 | 0.80 | 0.45 |
| TP9-TP10 | 0.25 | 0.10 | 0.30 | 0.07 | -1.52 | 0.16 | 0.34 | 0.07 | 0.34 | 0.08 | 0.07 | 0.95 |
| Gamma PLV |  |  |  |  |  |  |  |  |  |  |  |  |
| AF7-AF8 | 0.41 | 0.29 | 0.36 | 0.26 | 0.44 | 0.67 | **0.28** | **0.11** | **0.18** | **0.06** | **3.17** | **0.02** |
| AF7-TP9 | 0.49 | 0.33 | 0.41 | 0.30 | 0.58 | 0.58 | 0.24 | 0.09 | 0.23 | 0.15 | 0.15 | 0.89 |
| AF7-TP10 | 0.45 | 0.31 | 0.33 | 0.26 | 0.89 | 0.39 | 0.24 | 0.10 | 0.26 | 0.21 | -0.32 | 0.76 |
| AF8-TP9 | 0.56 | 0.35 | 0.53 | 0.33 | 0.28 | 0.78 | 0.29 | 0.28 | 0.19 | 0.16 | 0.81 | 0.44 |
| AF8-TP10 | 0.53 | 0.36 | 0.51 | 0.28 | 0.17 | 0.87 | 0.31 | 0.26 | 0.21 | 0.15 | 0.82 | 0.44 |
| TP9-TP10 | 0.91 | 0.14 | 0.82 | 0.23 | 2.26 | 0.05 | 0.76 | 0.32 | 0.62 | 0.26 | 1.01 | 0.35 |

*Note.* ^*^*p* < .05, ^**^*p* < .01, *p values are corrected for False Discovery Rate (FDR)*

**Supplementary Materials**

**Supplementary Table 4.**

Pearson’s Correlation between change in EEG metrics and proportional change in HAMD score across all subjects

| Variable | Correlation Coefficient | *p* | Correlation Coefficient _Active Group | *p*_Active Group | Correlation Coefficient _Sham Group | *p*_Sham Group |
| --- | --- | --- | --- | --- | --- | --- |
| Full Power |  |  |  |  |  |  |
| AF7 | 0.25 | 0.31 | -0.16 | 0.7 | 0.36 | 0.27 |
| AF8 | 0.22 | 0.37 | -0.18 | 0.66 | -0.34 | 0.31 |
| TP9 | 0.2 | 0.43 | 0.71 | 0.05 | -0.18 | 0.59 |
| TP10 | -0.41 | 0.09 | -0.58 | 0.13 | -0.26 | 0.45 |
| Delta Power |  |  |  |  |  |  |
| AF7 | 0.34 | 0.18 | 0.45 | 0.26 | 0.09 | 0.8 |
| AF8 | 0.2 | 0.43 | -0.77 | 0.02 | 0.35 | 0.29 |
| TP9 | 0.04 | 0.9 | -0.17 | 0.69 | 0.32 | 0.34 |
| TP10 | -0.1 | 0.71 | 0.34 | 0.41 | 0.02 | 0.95 |
| Theta Power |  |  |  |  |  |  |
| AF7 | -0.01 | 0.98 | 0.62 | 0.1 | -0.12 | 0.72 |
| AF8 | 0.21 | 0.43 | 0.24 | 0.56 | -0.15 | 0.66 |
| TP9 | 0.27 | 0.31 | -0.15 | 0.72 | 0.32 | 0.34 |
| TP10 | 0.06 | 0.83 | -0.31 | 0.45 | 0.48 | 0.13 |
| Alpha Power |  |  |  |  |  |  |
| AF7 | 0.34 | 0.2 | 0.53 | 0.18 | 0.06 | 0.85 |
| AF8 | -0.21 | 0.46 | -0.6 | 0.11 | -0.06 | 0.87 |
| TP9 | 0.31 | 0.26 | 0.21 | 0.61 | -0.02 | 0.95 |
| TP10 | 0.17 | 0.55 | -0.19 | 0.65 | 0.18 | 0.6 |
| Beta Power |  |  |  |  |  |  |
| AF7 | 0.15 | 0.58 | -0.34 | 0.41 | 0.12 | 0.72 |
| AF8 | 0.03 | 0.92 | -0.04 | 0.92 | 0.03 | 0.93 |
| TP9 | 0.03 | 0.91 | -0.08 | 0.85 | 0.15 | 0.65 |
| TP10 | 0.07 | 0.81 | 0.44 | 0.27 | 0.1 | 0.77 |
| Gamma Power |  |  |  |  |  |  |
| AF7 | -0.17 | 0.57 | -0.16 | 0.71 | 0.29 | 0.39 |
| AF8 | -0.39 | 0.18 | -0.31 | 0.45 | 0.19 | 0.57 |
| TP9 | 0.03 | 0.92 | -0.33 | 0.42 | -0.05 | 0.88 |
| TP10 | -0.32 | 0.28 | -0.47 | 0.24 | -0.06 | 0.86 |
| Full PLV |  |  |  |  |  |  |
| AF7, AF8 | 0.27 | 0.27 | 0.73 | 0.04 | 0.05 | 0.88 |
| AF7, TP9 | 0.16 | 0.53 | 0.15 | 0.71 | -0.01 | 0.98 |
| AF7, TP10 | 0.24 | 0.31 | 0.63 | 0.1 | 0.09 | 0.8 |
| AF8, TP9 | 0.24 | 0.33 | 0.74 | 0.04 | 0.13 | 0.7 |
| AF8, TP10 | 0.41 | 0.08 | 0.81 | 0.02 | 0.28 | 0.4 |
| TP9, TP10 | 0.39 | 0.1 | 0.81 | 0.01 | 0.07 | 0.84 |
| Delta PLV |  |  |  |  |  |  |
| AF7, AF8 | 0.08 | 0.73 | -0.14 | 0.74 | 0.22 | 0.51 |
| AF7, TP9 | -0.09 | 0.71 | -0.15 | 0.73 | -0.25 | 0.46 |
| AF7, TP10 | -0.12 | 0.63 | -0.28 | 0.51 | -0.07 | 0.84 |
| AF8, TP9 | -0.09 | 0.71 | -0.2 | 0.64 | -0.09 | 0.79 |
| AF8, TP10 | 0.27 | 0.27 | 0.79 | 0.02 | 0 | 1 |
| TP9, TP10 | 0.13 | 0.59 | 0.72 | 0.04 | -0.22 | 0.52 |
| Theta PLV |  |  |  |  |  |  |
| AF7, AF8 | 0.23 | 0.34 | 0.79 | 0.02 | -0.04 | 0.91 |
| AF7, TP9 | 0.35 | 0.14 | 0.85 | 0.01 | 0.13 | 0.71 |
| AF7, TP10 | 0.37 | 0.12 | 0.86 | 0.01 | 0.16 | 0.65 |
| AF8, TP9 | 0.45 | 0.05 | 0.88 | 0 | 0.01 | 0.98 |
| AF8, TP10 | 0.02 | 0.94 | -0.18 | 0.66 | 0.23 | 0.5 |
| TP9, TP10 | -0.28 | 0.25 | 0 | 0.99 | -0.33 | 0.32 |
| Alpha PLV |  |  |  |  |  |  |
| AF7, AF8 | 0.23 | 0.34 | -0.4 | 0.32 | -0.04 | 0.91 |
| AF7, TP9 | 0.35 | 0.14 | 0.17 | 0.69 | -0.08 | 0.82 |
| AF7, TP10 | 0.37 | 0.12 | 0.62 | 0.1 | -0.17 | 0.62 |
| AF8, TP9 | 0.45 | 0.05 | 0.81 | 0.01 | -0.17 | 0.61 |
| AF8, TP10 | 0.02 | 0.94 | 0.19 | 0.66 | 0.3 | 0.37 |
| TP9, TP10 | -0.28 | 0.25 | 0.77 | 0.03 | 0.01 | 0.98 |
| Beta PLV |  |  |  |  |  |  |
| AF7, AF8 | -0.07 | 0.77 | 0.6 | 0.12 | -0.2 | 0.56 |
| AF7, TP9 | 0.4 | 0.09 | 0.88 | 0 | 0.18 | 0.6 |
| AF7, TP10 | 0.28 | 0.25 | -0.14 | 0.74 | 0.36 | 0.28 |
| AF8, TP9 | -0.36 | 0.13 | 0.43 | 0.29 | -0.35 | 0.29 |
| AF8, TP10 | -0.01 | 0.96 | 0.7 | 0.05 | -0.19 | 0.58 |
| TP9, TP10 | -0.36 | 0.13 | -0.58 | 0.13 | -0.26 | 0.44 |
| Gamma PLV |  |  |  |  |  |  |
| AF7, AF8 | -0.04 | 0.87 | 0.16 | 0.7 | -0.11 | 0.76 |
| AF7, TP9 | 0.03 | 0.89 | -0.47 | 0.24 | 0.16 | 0.63 |
| AF7, TP10 | -0.07 | 0.76 | -0.22 | 0.61 | 0.05 | 0.89 |
| AF8, TP9 | 0.11 | 0.66 | 0.23 | 0.58 | 0 | 0.99 |
| AF8, TP10 | 0.14 | 0.56 | 0.4 | 0.32 | -0.02 | 0.95 |
| TP9, TP10 | -0.08 | 0.73 | -0.11 | 0.79 | -0.25 | 0.47 |

*Note.* ^*^*p* < .05, ^**^*p* < .01, *p values are corrected for False Discovery Rate (FDR)*

**Supplementary Materials**

**Supplementary Table 5.**

The classification accuracies for different deep learning models for different EEG bands.

| **EEG frequency band with which PLV features extracted** | **1DCNN classification performance** | | | **Fully connected perceptron network performance** | | |
| --- | --- | --- | --- | --- | --- | --- |
|  | **Accuracy** | **Sensitivity** | **Specificity** | **Accuracy** | **Sensitivity** | **Specificity** |
| Full band | **46.48** | 14.52 | 65.12 | 44.95 | 19.38 | 59.87 |
| Delta band | **60** | 15.2 | 86.2 | 56.33 | 20.1 | 77.46 |
| Theta band | **67.5** | 36.1 | 85.7 | 66.17 | 50.9 | 75.06 |
| Alpha band | **68.42** | 14.2 | 100 | 56.94 | 17.54 | 79.93 |
| Beta band | 68.21 | 29.17 | 90.9 | **71.19** | 42.43 | 87.96 |
| Gamma band | **65.8** | 30.5 | 86.4 | 58.64 | 41.21 | 68.82 |

**Supplementary Materials**

**Supplementary Table 6.**

The classification accuracies for different deep learning models for the combined PLV from different EEG bands.

| **EEG frequency band combinations** | **1DCNN classification performance** | | | **Fully connected perceptron network performance** | | |
| --- | --- | --- | --- | --- | --- | --- |
|  | **Accuracy** | **Sensitivity** | **Specificity** | **Accuracy** | **Sensitivity** | **Specificity** |
| Delta, Theta | 63.8 | 31 | 83 | 65.85 | 56.29 | 71.43 |
| Delta, Alpha | 56.8 | 16.2 | 80.5 | 52.5 | 17.08 | 73.16 |
| Delta, Beta | 67.7 | 36.2 | 86.1 | 67.17 | 45.47 | 79.83 |
| Delta, Gamma | 62.77 | 40.6 | 75.7 | 61.72 | 45.74 | 71.04 |
| Theta, Alpha | 68.7 | 47.4 | 81.1 | 66.64 | 47.26 | 77.94 |
| Theta, Beta | **71.4** | 53.3 | 82 | 69.85 | 54.13 | 79.03 |
| Theta, Gamma | 59.59 | 39.39 | 71.37 | 59.81 | 47.13 | 67.2 |
| Alpha, Beta | 65.49 | 29.6 | 86.4 | 64.78 | 32.19 | 83.79 |
| Alpha, Gamma | 47.28 | 29.33 | 57.09 | 48.86 | 33.53 | 57.8 |
| Beta, Gamma | 58.05 | 42.42 | 67.17 | 47.58 | 32.07 | 56.63 |
| Delta, Theta, Alpha | 66.16 | 48.9 | 76.21 | 61.76 | 43.67 | 72.3 |
| Delta, Theta, Beta | 66.36 | 40 | 81.76 | 67.7 | 45.31 | 80.75 |
| Delta, Theta, Gamma | 65.1 | **59.3** | 68.5 | 60.76 | 52.06 | 65.84 |
| Delta, Alpha, Beta | 66.56 | 39.88 | 82.12 | 63.07 | 33.7 | 80.21 |
| Delta, Alpha, Gamma | 53.59 | 42.92 | 59.8 | 55.18 | 44.73 | 61.27 |
| Delta, Beta, Gamma | 57.21 | 41.23 | 66.54 | 60.34 | 45.64 | 68.91 |
| Theta, Alpha, Beta | **71.94** | 52.88 | 83.06 | 67.16 | 44.11 | 80.6 |
| Theta, Alpha, Gamma | 64.44 | 51.6 | 71.8 | 57.32 | 43.29 | 65.5 |
| Theta, Beta, Gamma | 60.47 | 49.52 | 66.82 | 58.93 | 45.02 | 67.05 |
| Alpha, Beta, Gamma | 59.16 | 47.08 | 66.21 | 51.13 | 43.76 | 55.43 |
| Delta, Theta, Alpha, Beta | 63.9 | 45.47 | 74.7 | **66.32** | 43.9 | 79.4 |
| Delta, Theta, Alpha, Gamma | 62.03 | 53.17 | 67.18 | 56.9 | 46.04 | 63.26 |
| Delta, Theta, Beta, Gamma | 61.68 | 56.82 | 64.52 | 60.68 | 48.52 | 67.78 |
| Delta, Alpha, Beta, Gamma | 56.9 | 44.9 | 63.87 | 54.15 | 37.72 | 63.73 |
| Theta, Alpha, Beta, Gamma | 60.15 | 54.44 | 63.48 | 54.5 | 45.17 | 59.95 |
| Delta, Theta, Alpha, Beta, Gamma | 62.25 | 54.48 | 66.78 | 56.95 | 44.46 | 64.24 |
